# ‘No lockdown’ policy for COVID-19 epidemic in Bangladesh : Good, bad or ugly?

**DOI:** 10.1101/2020.10.21.20216812

**Authors:** Zakaria Shams Siam, M. Arifuzzaman, Md. Harunur Rashid, Md. Shariful Islam

## Abstract

Bangladesh has been combating the COVID-19 pandemic with limited financial resources and poor health infrastructure since March, 2020. Although the government has imposed several restricted measures to curb the progression of the outbreak, these arrays of measures are not sustainable in the long run. In this study, we assess the impact of lift of flexible lockdown on the COVID-19 dynamics in Bangladesh. Our analysis demonstrates that the country might experience second infection peak in 6-7 months after the withdrawal of current lockdown. Moreover, a prolonged restrictions until January, 2021 will shift the infection peak towards August, 2021 and will reduce approximately 20 % COVID-19 cases in Bangladesh.

**What we know:**

- Bangladesh has been going through COVID-19 crisis and in response, the Government has implemented restricted array of measures to curb the COVID-19 outbreak in Bangladesh.

**What this article adds:**

- The impact of ‘no lockdown’ policies on COVID-19 pandemic in Bangladesh.
- Appearance of second infection peak in 6-7 months after the withdrawal of current lockdown.

## Introduction

COVID-19 has emerged as one of the most challenging infectious diseases in the World’s history. ^1,2^ Initially, China, several countries of Europe and America were severely affected by COVID-19, but recently the virus has spread in South-Asian countries including, India, Pakistan, and Bangladesh. Although several studies demonstrate the importance of imposing lockdown to slow down the progression of the pandemic ^3,4^, a lockdown cannot last indefinitely as it has a negative impact on national economies as well as people’s goodwill. As a result, despite the significant amount of COVID-19 positive cases under restricted conditions, Government has lifted the lockdown gradually by reopening all the industries, markets and offices with a notable exception of educational institutes. Notably, the age group (in years) of 1-20 carries 43.5 % of total age distribution of the population in Bangladesh (collected from Socioeconomic Data and Application Center, or SEDAC). Moreover, Hoque and colleagues recently reported that only 10.2 % of the total COVID-19 positive cases were identified within the age group of 1-20 compared 54.6 % cases from the age group of 21-40.^5^ Since all the academic institutes are closed to contain COVID-19 diffusion, one might ask whether Bangladesh is going through a flexible lockdown and if so, then what could be the consequences on the public health when the flexible lockdown is lifted in the upcoming months. Besides, it is important to have a trade-off between various key epidemiological factors to predict the COVID-19 transmission in Bangladesh and that is where a mathematical estimation becomes relevant to evaluate the future we may expect based on the latest policies implemented in the country. Here, we use the epidemiological model to explore the consequences of ‘No lockdown’ policy on the future trend of COVID-19 cases in Bangladesh so that policymaker can take right decisions considering the socioeconomic structure of the country.

## Methods

SEIRD model is used to forecast the dynamics of the pandemic in Bangladesh. The model has five time dependent important variables: *S, E, I, R*, and *D*.^6^ The terms *S(t), E(t), I(t), R(t) and D(t)* denote the susceptible population, exposed people, infected population, recovered people and the death individuals respectively.

The total population at time *t*: *N*(*t*) = *S*(*t*) + *E*(*t*) + *I*(*t*) + *R*(*t*) + *D*(*t*).

A set of ordinary differential equations is used (see the supplement) to describe the SEIRD model as shown previously.^6^ The basic reproduction number (R_0_) plays a key role to identify whether an infectious disease becomes epidemic (R_0_ > 1) or not (R_0_ < 1) in any epidemiological analyses. Therefore, the epidemic control methods are executed to stop the outbreak when R_0_ > 1.^7^ The time-dependent reproduction number, R_t_, determines the transmission speed of the pandemic. In this study, we set the value of basic reproduction number, R_0_ ∼2.5.

A set of differential equations that describes the SEIRD model

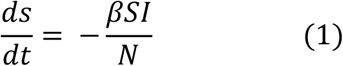

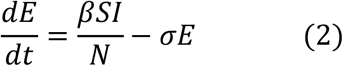

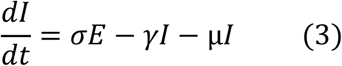

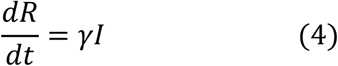

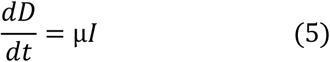

The parameters *β, γ* and µ represent the transmission rate, recovery rate and death rate respectively. The symbol σ stands for the fraction of the rate of transfer of disease from latent to the infectious stage.

### Parameter values

In our study, we selected the following initial conditions: E(0) = 10, I(0) = 40, R(0) = 0 and D(0) = 0. To take the correct susceptible population is very important to model any pandemic where the entire population can be chosen as susceptible population. Several studies reported an estimation of susceptible by multiplying 10^−3^ - 10^−5^ with the total population of the country based on Germany, South Korea and China’s COVID-19 outbreak.^4^ In this note, S(0) ∼ N ^*^ 10^−1^ (17 million) has been taken as it provides the best fit with real data. Moreover, E(0) = 10 was set in the analysis as it would allow the disease to spread at a moderate rate. We tested several values of E(0) (not shown in the manuscript) and the value of 10 gives the best fit. The value of transmission rate, *β* = 0.12 was set for the analysis, whereas recovery rate, γ = 0.038 day^-1^ was considered which corresponds to ∼ 26 days. The value of death rate, µ, was taken as 0.0008.

## Results

Imposing restricted array of measures can have positive impacts on the COVID-19 outbreak,^3,4^ but lockdown cannot last forever without causing enormous damage to national economies and compromising people’s resilience. In this note, we forecast the infection spread and death tolls, if the current confinement is withdrawn before COVID-19 pandemic ends in Bangladesh. The GNU octave (Version 5.2.0) and Maple have been used for the SEIRD model simulation and plotting the data. We consider the functional form of time-dependent reproduction number, R_t_, and demonstrate how the lifting of the nationwide confinement at different times might remodel the location of infection and death peak. The real data keep the R_t_ values in the range 0.96 ≤ R_t_ ≤ 3.8 at the beginning of the pandemic similar to other reports,^3,8^ whereas we assume a gradual increase (0.96 ≤ R_t_ ≤ 1.3) of R_t_ values once the restricted measures are completely taken off in the upcoming months. Thus, based on the current situation, if the lockdown is lifted in Bangladesh starting from 1^st^ of October, 2020, our model predicts the occurrence of next infection peak (∼400000 active cases) at the beginning of June (5^th^-9^th^ June), 2021 (Fig. 1A), with a daily death toll going as high as 200 deaths (Fig. 1B). Moreover, Bangladesh might face the dire consequences by then as approximately 85000 citizens could die due to the COVID-19 outbreak. We further explore the probable fate of coronavirus affected Bangladesh if the nation exits lockdown from November, 2020. In that case, the present epidemiological model forecasts the reappearance of infection peak (∼370000 active cases) in the third week of July, 2021 with ∼75000 deaths associated with coronavirus. Notably, prolonging the current flexible lockdown until January, 2021 or withdrawing the restrictions in January, 2021, will further shift the infection peak to the middle of September, 2021 (Fig. 1A). In that case, the SEIRD model predicts a total infected number ∼330000 and death toll of 70000 in the third week of September, 2021 (Fig. 1B). Therefore, this study exclusively preditcs COVID-19 pandemic situation in Bangladesh, given the potential for lift of lockdown at different time points before the outbreak ends.

**Figure 1.**
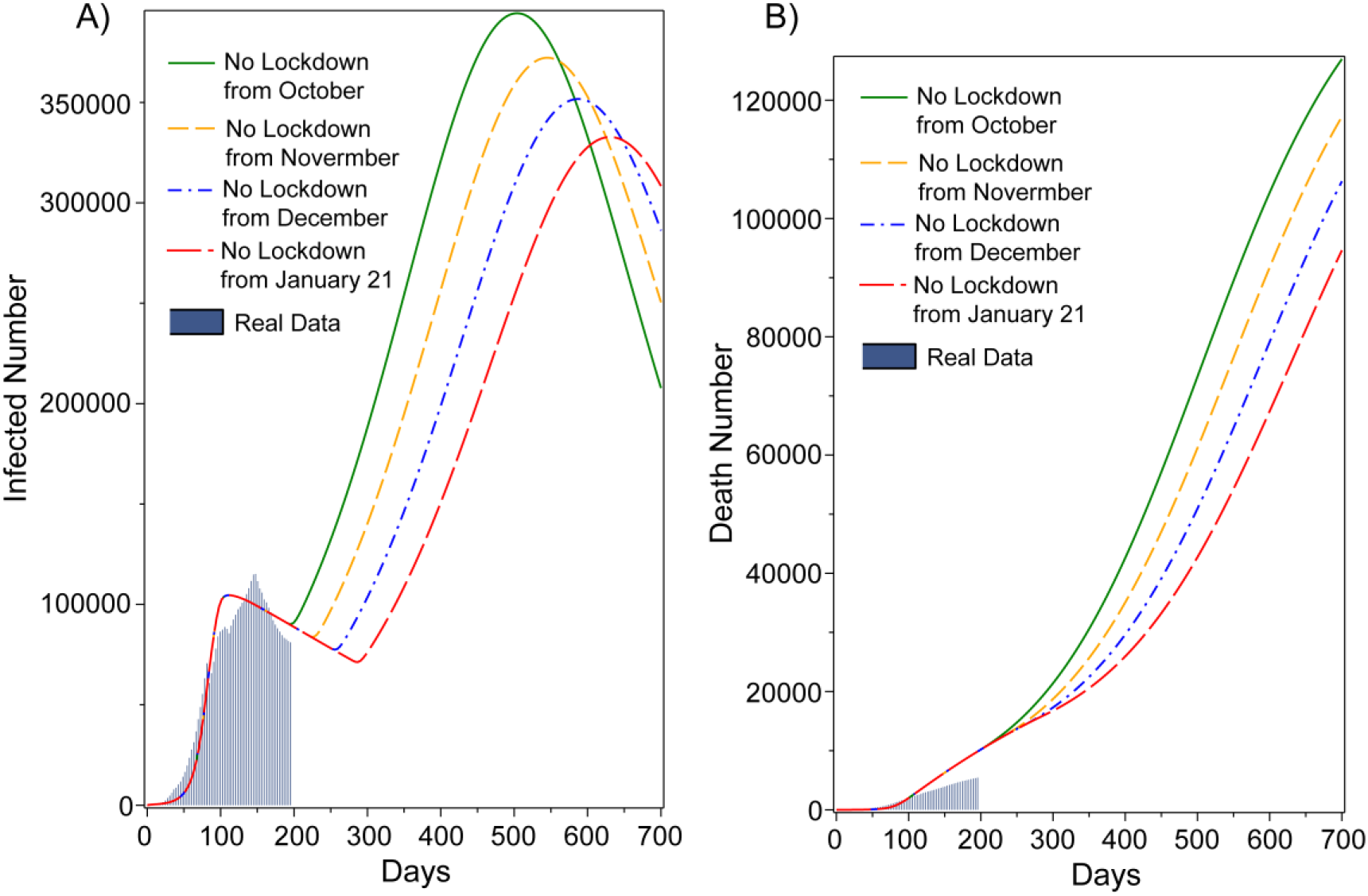
A-B) Considering a gradual increase (0.96 ≤ R_t_ ≤ 1.3) of R_t_, dynamics of the infected population and death tolls as a function of time in Bangladesh, when the lockdown is lifted from October, 2020 to January, 2021. Withdrawing lockdown on 1^st^ October, 2020 clearly creates the appearance of second infection peak in June, 2021. The real data are fitted from March 26 to October 10, 2020.

**Figure 2.**
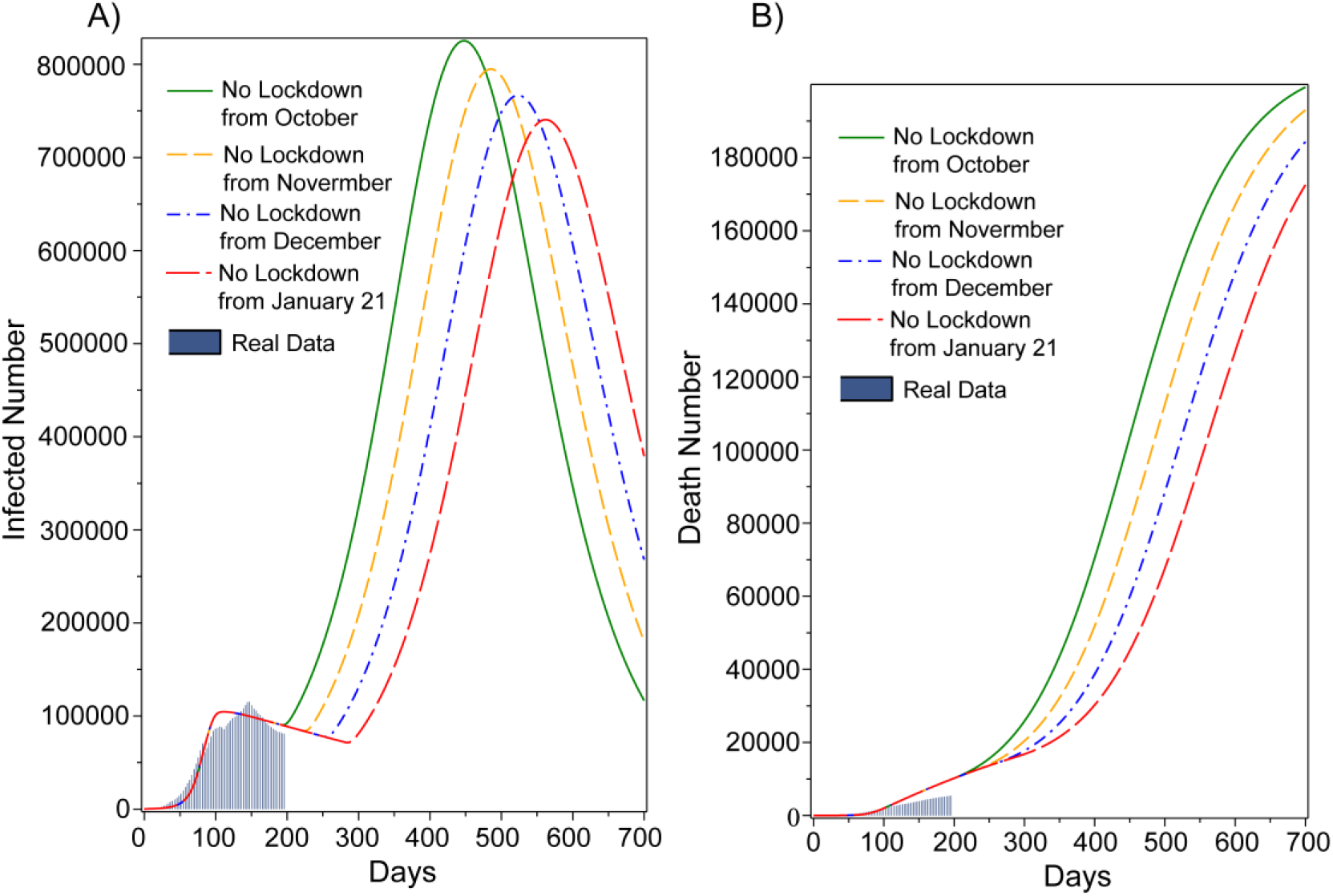
A-B) Considering a gradual increase (0.96 ≤ R_t_ ≤ 1.5) of R_t_, dynamics of the infected population and death tolls as a function of time in Bangladesh, when the lockdown is lifted from October, 2020 to January, 2021. Withdrawing lockdown on 1^st^ October, 2020 clearly creates the appearance of second infection peak in April, 2021. The real data are fitted from March 26 to October 10, 2020.

To estimate the dire consequences due to the current pandemic, next we assume a gradual increase the R_t_ values from 0.96 to 1.5 in no restricted conditions. Similar to our first study, no lockdown from October, 2020 results the next infection peak with ∼820000 corona positive cases on May, 2021. Moreover, if the Government extend the flexible lockdown until January, 2021, the infection peak will touch approximately 750000 cases along with ∼110000 deaths on August, 2021. Moreover, we also make further speculation based on the R_t_ values (0.96 ≤ R_t_ ≤ 2) in no lockdown environment (Supplement Fig. 1) in Bangladesh.

## Discussion & Conclusion

Several reports showed the major role of lockdown to slow down the spread of COVID-19 and in response, the Bangladesh government has set up various strict measures to keep the rising of outbreak under control.^6,8^ However, as a middle-income country, Bangladesh might not sustain being restricted in the long run by imposing prolonged lockdown. Hence, in this study, we forecast the expected consequences of the lift of lockdown and show ‘no lockdown’ policies could be jeopardized as it skyrockets the number of COVID-19 positive cases and the death toll in Bangladesh. Our epidemiological model further predicts about a gigantic infection peak with enormous death tolls (∼75000 deaths) that might start appearing approximately 6-7 months after the withdrawal of current lockdown. Strikingly, we notice that a prolonged lockdown until January, 2021 might reduce 20 % COVID-19 infected cases on September, 2021. In addition, this policy will open a window of 11 months to rebuild the health infrastructure in Bangladesh. Moreover, it will also bring some hope of getting the novel COVID-19 vaccine to defeat the coronavirus outbreak. As Bangladesh encounters a three-way tug of war, a battle between the coronavirus pandemic, reviving the national economy and keeping society on an even keel, the policymakers need to come up with more appropriate lockdown exit framework in order to keep a sustainable socioeconomic structure of Bangladesh. In summary, we hope that our study will be useable for the policy speculation and regulating the future outbreaks in Bangladesh.

## Data Availability

All the data are available with the manuscript

## Acknowledgement

The authors would like to thank Dr. Fariha Jasmin Chowdhury for their critical comments as a general reader.

## Author’s contributions

MA, ZS and MI made substantial contributions to the conception or design of the work. ZS and MA completed the relevant studies and created plots related to the SEIRD estimation, while MA, MH and MI wrote the draft and critically revised the manuscript. All the authors met regularly to discuss the outcome of each experiment. The data were collected from the IEDCR website and each of the results was cross checked by all authors. All authors are responsible for acquiring, analyzing, and interpreting the data for this article.

## Availability of data and materials

All data were collected from the IEDCR, Bangladesh website.

## Supplement

**Supplement figure 1.**
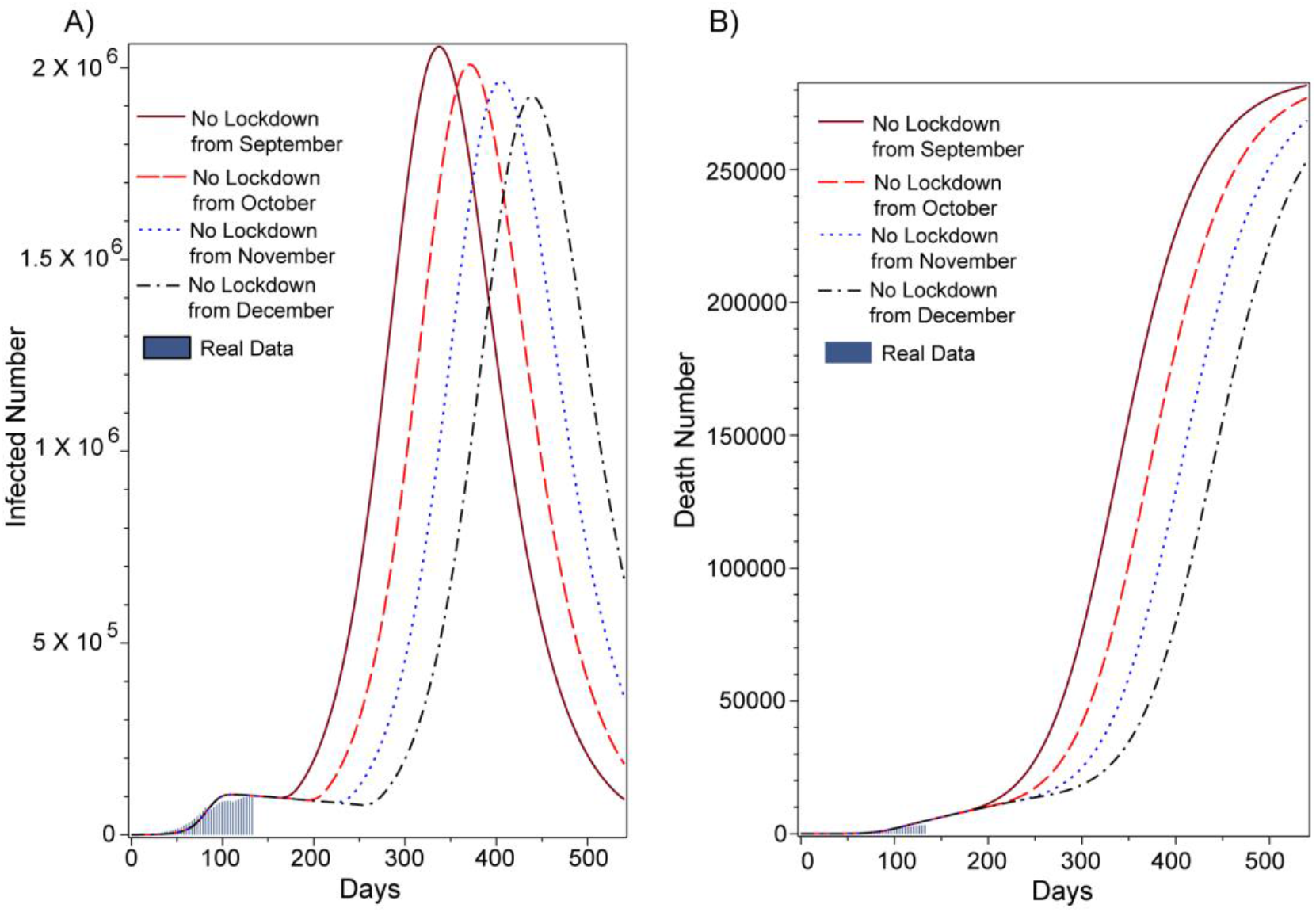
A-B) Considering a gradual increase (0.96 ≤ R_t_ ≤ 2) of R_t_, dynamics of the infected population and death tolls as a function of time in Bangladesh, when the lockdown is lifted from September, 2020 to December, 2020. Withdrawing lockdown on 1^st^ October, 2020 clearly creates the appearance of second infection peak in April, 2021. The real data are fitted from March 26^th^ to October 10^th^, 2020.

